# Evaluating Traditional, Machine Learning, and Deep Learning Models for Predicting Outpatient Healthcare Utilization in Europe: A Longitudinal Panel Analysis

**DOI:** 10.1101/2025.09.09.25335390

**Authors:** Vincent Cheng-Sheng Li, Tzu-Pin Lu, Charlotte Wang, Kanya Anindya, John Tayu Lee

## Abstract

Accurate prediction of outpatient utilization supports responsive health-care planning. Using longitudinal micro-data from the Survey of Health, Ageing and Retirement in Europe (SHARE)—10,777 adults observed across Waves 5, 6, 8, and 9 (2013, 2015, 2019–2020, and 2021–2022, respectively)—we forecast Wave-9 outpatient visit counts from Waves 5–6–8 history. We benchmark generalized estimating equations (GEE) and generalized linear mixed models (GLMM) against sequence-aware neural architectures—LSTM, GRU, CNN, CNN-LSTM, CNN-GRU, and a feed-forward DNN—under strict leakage guards (no Wave-9 predictors), train-only imputation/scaling, and a fixed 80/20 row-level split with performance uncertainty estimated via 400 bootstrap replicates. Baselines follow standard GEE/GLMM formulations.

Across models, adding a second historical wave materially improves accuracy, with smaller gains from a third wave. The best configuration—a CNN–GRU with a shallow causal 1-D convolutional front-end and a two-layer GRU head—achieves MAE = 3.41 (95% CI 3.24–3.59) and RMSE = 4.91 (4.60–5.23) on the held-out test set, modestly outperforming a tuned GRU (MAE = 3.46; RMSE = 5.21) and the other deep baselines. Overall, results indicate that sequence models capture short-horizon temporal structure in SHARE, but incremental gains hinge more on temporal depth and careful regularization than on architectural complexity alone. Methodologically, the study demonstrates a leak-safe pipeline for panel forecasting in population health using modern recurrent/convolutional networks alongside classical longitudinal benchmarks.

## INTRODUCTION

Accurately predicting health service utilization is a critical component of effective health policy formulation and healthcare system planning ^1^. In the face of rising pressures from aging populations, the increasing prevalence of chronic diseases, and constrained resources, European health systems must anticipate demand for services—including primary care, hospitalizations, and long-term care—to ensure sustainability and efficiency ^2^. Reliable forecasts enable targeted interventions, inform strategic resource allocation, and support evidence-based policy, ultimately improving population health and economic resilience.

Traditional empirical approaches to modeling healthcare use predominantly rely on parametric regression frameworks, such as generalized linear models (GLMs) and mixed-effects models ^3^. These methods offer tractable inference, transparent assumptions, and interpretable parameters, making them particularly suitable for policy evaluation and causal analysis ^4^. However, the structural assumptions underpinning these models, including linearity and additive separability, may limit their ability to capture complex, non-linear relationships and high-order interactions among the diverse set of covariates commonly observed in health data ^5^. Moreover, in longitudinal settings, the presence of dynamic dependencies and unobserved heterogeneity further challenges the predictive performance of conventional techniques ^6^.

Recent methodological advances in machine learning (ML), including neural networks (NNs), offer alternative strategies for prediction that are less reliant on restrictive parametric assumptions. These models are designed to approximate complex, potentially non-linear data-generating processes and may therefore yield superior predictive accuracy, particularly in the presence of large, high-dimensional, and longitudinal datasets ^7^. Despite their increasing prominence in adjacent fields, the application of ML techniques to healthcare utilization prediction remains limited, and their relative performance vis-à-vis traditional econometric models has not been systematically assessed in this context ^8^.

This study contributes to the literature by conducting a rigorous comparative evaluation of three modeling paradigms—traditional regression models, general-purpose machine learning algorithms, and neural networks—in the prediction of healthcare service utilization. Leveraging harmonized, multi-wave panel data from multiple European countries, we assess the out-of-sample predictive performance of each approach under varying temporal data conditions and across multiple types of service outcomes.

Specifically, we examine four hypotheses: (1) machine learning models will achieve higher predictive accuracy than traditional regression models in forecasting future healthcare use. (2) The incorporation of additional longitudinal waves will improve predictive accuracy across all modeling approaches, with more pronounced gains for machine learning models due to their ability to exploit temporal structures. (3) The relative importance of covariates will be broadly similar across models, suggesting robustness of predictor relevance despite methodological differences. (4) Under limited data scenarios (e.g., single or two-wave panels), the performance gap between traditional regression models and more complex machine learning models will be attenuated.

## METHODS

### Data source and cohort

Data were drawn from the Survey of Health, Ageing and Retirement in Europe (SHARE), release 9.0.0, which provides harmonized panel data on adults aged 50 years and older across Europe. Respondents were eligible for inclusion if they participated in all of the following waves: Wave 5 (2013), Wave 6 (2015), Wave 8 (2019/20) and Wave 9 (2021/22). Individuals who participated only in the life-history-focused Wave 7 or in waves prior to Wave 5 were excluded. In addition, records missing a valid household identifier across all four target waves were removed. After applying these criteria, the final analytic cohort consisted of 11,972 individuals from 16 countries, each contributing data from three exposure waves and one outcome wave.

### Outcome definition

The primary outcome was the self-reported number of outpatient physician visits during the 12 months before the Wave-9 interview (2021/2022). To mitigate the influence of extreme values, top 1 percent of reported counts were seen as outliers and thus were removed. This variable serves as a proxy for the frequency of healthcare utilization in their life.

### Predictor variables

Candidate predictors were selected a priori from SHARE domains known to influence health-care utilization. Time-invariant covariates included sex, year of birth, country of residence, and educational attainment, all measured at Wave 5, which served as the baseline reference.

Time-varying covariates—extracted from Waves 5, 6, and 8—included prior outpatient and inpatient healthcare utilization, self-rated health, chronic disease indicators, body mass index (BMI), limitations in activities of daily living (ADLs), smoking status, frequency of alcohol consumption, physical activity, household composition, and household income quintile.

### Data processing

A series of preprocessing steps was applied to prepare the dataset for analysis.

First, variable types were reviewed to determine the appropriate transformation strategies. Binary categorical variables (exactly two levels) were represented as 0/1 indicators. Nominal variables with more than two unordered levels, and ordinal variables for which category spacing could not be assumed equal, were one-hot encoded using K−1 dummy variables to avoid imposing an artificial order and to prevent multicollinearity. The country of residence was represented using a set of dummy variables. Variables that were already numeric were retained without modification.

Next, variable names were aligned across survey waves based on their suffixes (e.g., _w5, _w6, _w8, _w9). To ensure consistency over time, only variables that were present in all three exposure waves were retained, resulting in a uniform feature set across time points.

To address missing data, columns with more than 10% missing values across the full sample were removed. Subsequently, any rows containing missing or infinite values were excluded. To reduce the influence of outliers, outpatient visit counts exceeding the 99th percentile were excluded.

Finally, the cleaned dataset was randomly split into training (80%) and test (20%) subsets. Continuous predictors were z-standardized using the mean and standard deviation from the training set; these parameters were then applied unchanged to the test set.

### Statistical baselines

As statistical baselines, we estimated two Poisson regression models: (i) a generalised estimating equation (GEE) with exchangeable correlation and (ii) a generalised linear mixed model (GLMM) with a random intercept for the subject.

Prior to model fitting, preprocessing steps were performed to ensure model stability and interpretability.. A Poisson LASSO regression was used for variable selection. Any remaining rank deficiency in the design matrix was addressed using QR decomposition.

All statistical models were implemented in R version 4.3.

### Deep Learning Models

We model longitudinal utilization as a three-step sequence using observations from

Waves 5, 6, and 8 to predict Wave 9. For each participant, dynamic features (those recorded at each wave) are stacked into a length-3 sequence; strictly non-wave columns form a static covariate vector. Leak guards exclude any predictor with a Wave 9 suffix, and all preprocessing statistics (imputation and scaling) are fit on the training split only and then applied to the held-out test set.

We evaluate six neural architectures: LSTM, GRU, DNN, CNN, and two hybrids— CNN–LSTM and CNN–GRU. Recurrent backbones use standard Keras LSTM/GRU layers; stacked variants set “return_sequences=True” in the first layer so the second layer consumes the hidden-state sequence. Hybrid models attach a shallow causal 1-D convolutional front end (Conv1D with optional dilation/stride) that transforms the 3×F sequence before passing it to the recurrent head (Figure 3). The CNN–GRU and CNN–LSTM reuse the best recurrent backbones (GRU and LSTM, respectively) discovered during backbone tuning; the convolutional front end is learned while keeping those backbone hyperparameters fixed. Training uses minibatches with early stopping on validation loss to prevent overfitting.

### Hyper-parameter tuning and sensitivity analyses

Model selection follows a two-stage protocol. First, we run Bayesian optimization with KerasTuner to identify promising regions of the search space (hidden units, dropout, learning rate for all models; plus the convolutional front-end for hybrids: number of Conv1D layers, filters, kernel size, dilation). Second, we conduct a focused grid search around the top Bayesian incumbents to finalize each architecture. Training uses early stopping on validation loss.

Because CNN-GRU yielded the strongest performance in the Waves 5–6–8 → Wave 9 task, we performed targeted ablations corresponding to Table 4 while keeping the GRU head fixed: (4a) optimizer×learning-rate sweeps (SGD, Adam); (4b) depth×filter-schedule (flat vs. tapered widths); (4c) kernel×depth with stride = 1 and causal padding; and (4d) regularization ablations toggling SpatialDropout1D, LayerNorm, and residual skips in the convolutional stack. These studies isolate which front-end design and training knobs most affect accuracy while holding the recurrent head constant.

### Evaluation and uncertainty

Model performance was evaluated on a held-out test set comprising 20% of the sample, with splits performed at the respondent level to ensure that all observations from the same individual were assigned to a single partition. Predictive accuracy was assessed using two primary metrics: mean absolute error (MAE) and root mean squared error (RMSE). These were selected to reflect both average and variance-weighted prediction error in the context of overdispersed count data.

To quantify uncertainty in model performance, we computed 95% confidence intervals for each metric using 400 bootstrap resamples of the test set.

### Model explainability

To place all models on comparable interpretability footing, we conducted a unified post-hoc analysis using SHAP (SHapley Additive exPlanations). For neural networks we used GradientExplainer when gradients were available and KernelExplainer otherwise. For each model we drew a background set of 200 observations and, where applicable, limited KernelExplainer to 1,000 evaluations. Global importance was summarized as the mean absolute SHAP value per feature, and we visualized the top ten predictors via violin (distribution) and bar (mean|SHAP|) plots; full SHAP matrices were archived for reproducibility.

To place the parametric models beside the SHAP summaries, we condensed the long coefficient lists from GEE and GLMM by grouping related terms (e.g., all levels of a multi-category item or the set of country dummies). We then ranked groups by a joint Wald magnitude that aggregates the usual Wald tests across the coefficients in each group; larger values indicate stronger overall evidence that the feature family contributes to the outcome. We display the top ten groups for each model to align with the SHAP top-10 convention.

### Software

*All analyses* were carried out in R and Python 3.12 with pandas 2.2, and *TensorFlow/Keras* 2.16 for deep-learning architectures. Cleaning routines were executed in Stata/SE 18.0. Source code is available at [github link].

### Ethical Considerations

The study was approved by the NTU Ethical Review Board (NTU REC-No.: 202504HM010) and used de-identified secondary data released by SHARE under its standard user agreement.

## RESULTS

### Participant characteristics

The analytic sample included 10,777 adults; ∼56% were women (Table 1). Chronic conditions were common: diabetes ∼29%, hypertension ∼18%, and cancer ∼51%; Parkinson’s disease was rare (∼0.8%). By country, Germany contributed the largest share (∼14%), followed by France (∼10%), Denmark (∼10%), and Switzerland (∼8%); Israel accounted for <1%.

**Table 1.** Descriptive summary of participants and outcome.

| Groups | Overall | Average Outpatient Count(%) |  |  |  |
| --- | --- | --- | --- | --- | --- |
|  |  | Wave 5 | Wave 6 | Wave 8 | Wave 9 |
|  | n (%) | n (%) | n (%) | n (%) | n (%) |
| **Gender** |  |  |  |  |  |
| male | 4,727<br>(43.86) | 5.63 (8.34) | 5.46 (6.86) | 6.42 (8.02) | 6.04 (5.90) |
| female | 6,050<br>(56.14) | 6.09 (8.25) | 5.64 (6.59) | 6.59 (8.27) | 5.99 (5.75) |
| **Diseases** |  |  |  |  |  |
| Heart | 1,108<br>(10.28) | 7.50 (9.61) | 7.20 (7.98) | 8.48 (10.81) | 7.80 (6.43) |
| Diabetes | 3,163<br>(29.35) | 7.45 (9.11) | 7.33 (7.71) | 8.61 (9.73) | 7.78 (6.58) |
| Parkinsons | 85<br>(0.79) | 10.21<br>(15.90) | 9.01 (8.44) | 11.51<br>(12.03) | 10.06 (8.35) |
| Hypertension | 1,899<br>(17.62) | 7.59 (9.05) | 7.73 (8.23) | 8.71 (9.67) | 7.78 (6.52) |
| Lung | 914<br>(8.48) | 8.39 (10.74) | 7.91 (8.30) | 9.07 (9.69) | 8.39 (7.28) |
| Cholesterol | 1,784<br>(16.55) | 7.52 (10.31) | 7.13 (8.23) | 8.22 (10.09) | 7.99 (6.81) |
| Cancer | 5,513<br>(51.16) | 7.15 (9.37) | 6.63 (7.37) | 7.68 (8.88) | 6.95 (6.24) |
| Stroke | 1,335<br>(12.39) | 8.39 (10.73) | 8.20 (8.80) | 9.37 (10.72) | 8.13 (7.27) |
| Ulcer | 3,825<br>(35.49) | 6.85 (8.84) | 6.57 (7.03) | 7.60 (8.68) | 7.03 (6.12) |
| **Countries** |  |  |  |  |  |
| Denmark | 1,072<br>(9.95) | 4.55 (6.90) | 4.05 (4.80) | 5.18 (8.03) | 4.52 (5.07) |
| Spain | 285<br>(2.64) | 6.02 (6.32) | 5.22 (5.64) | 6.97 (9.48) | 4.95 (5.43) |
| Italy | 656<br>(6.09) | 6.32 (8.61) | 6.26 (7.74) | 7.48 (7.73) | 6.34 (6.09) |
| Switzerland | 869<br>(8.06) | 5.12 (8.77) | 4.13 (5.03) | 5.10 (7.17) | 4.90 (5.29) |
| Slovenia | 666<br>(6.18) | 4.68 (6.09) | 4.88 (5.95) | 5.78 (6.14) | 4.71 (4.81) |
| France | 1,032<br>(9.58) | 5.85 (8.42) | 4.89 (4.65) | 5.77 (6.20) | 5.42 (4.59) |
| Israel | 64<br>(0.59) | 6.47 (8.87) | 4.48 (3.31) | 6.98 (12.51) | 5.88 (4.26) |
| Germany | 1,490<br>(13.83) | 7.85 (10.34) | 6.86 (7.71) | 7.77 (8.84) | 7.82 (6.61) |
| Belgium | 843<br>(7.82) | 7.21 (10.45) | 6.85 (8.18) | 7.98 (9.62) | 7.47 (6.72) |

Average outpatient utilization increased from Wave 5 to Wave 8 and then declined modestly in Wave 9. Among men, the mean number of visits was 5.63, 5.46, 6.42, and 6.04 across Waves 5, 6, 8, and 9, respectively; women showed a similar pattern (6.09, 5.64, 6.59, 5.99). Utilization was consistently higher among participants with chronic conditions. For example, those with diabetes averaged ∼7.3–8.6 visits across waves, and participants with stroke or lung disease averaged ∼8–9 visits in Wave 9, compared with ∼6 visits overall. Cross-country variation persisted, with Wave-9 means ranging from 4.52 visits in Denmark to 7.82 in Germany.

### Overall predictive performance

Table 2 (and Figure 1) reports out-of-sample accuracy for models trained on Waves 5–6–8 and evaluated on Wave 9. Mixed-effects models provide reasonable baselines: GLMM (MAE 3.749 (3.656–3.846); RMSE 5.627 (5.411–5.860)) improves upon GEE (MAE 4.003 (3.912–4.100); RMSE 5.851 (5.603–6.166)). All neural architectures outperform these baselines. Among sequence models, CNN-GRU attains the best overall accuracy (MAE 3.41 (3.24–3.59); RMSE 4.910 (4.60–5.23)), offering a modest but consistent improvement over the GRU baseline (MAE 3.46 (3.30–3.63); RMSE 5.21 (4.87–5.58)). Other neural configurations cluster closely: CNN-LSTM (MAE 3.51 (3.35–3.69); RMSE 5.29 (4.96–5.67)), CNN (MAE 3.52 (3.37–3.70); RMSE 5.260 (4.95– 5.61)), DNN (MAE 3.49 (3.34–3.68); RMSE 5.300 (4.98–5.67)), and LSTM (MAE 3.60 (3.44–3.79); RMSE 5.34 (5.01–5.71)). In summary, non-linear sequence models yield the highest fidelity for Wave-9 outpatient prediction, with CNN-GRU leading and GRU a close second; gains across neural variants are incremental yet robust relative to mixed-effects baselines. We report MAE and RMSE with 95% bootstrap confidence intervals following standard regression-evaluation practice and ACM manuscript conventions.

**Table 2.** Overall Models Performances using Waves 5, 6, 8 to Predict Wave 9.

| **Model** | **MAE (95% CI)** | **RMSE (95% CI)** |
| --- | --- | --- |
| GEE | 4.003 (3.912–4.100) | 5.851 (5.603–6.166) |
| GLMM | 3.749 (3.656–3.846) | 5.627 (5.411–5.860) |
| LSTM | 3.60 (3.44–3.79) | 5.34 (5.01–5.71) |
| CNN-LSTM | 3.51 (3.35–3.69) | 5.29 (4.96–5.67) |
| CNN-GRU | 3.41 (3.24–3.59) | 4.91 (4.60–5.23) |
| CNN | 3.52 (3.37–3.70) | 5.26 (4.95–5.61) |
| GRU | 3.46 (3.30–3.63) | 5.21 (4.87–5.58) |
| DNN | 3.49 (3.34–3.68) | 5.30 (4.98–5.67) |

**Figure 1.**
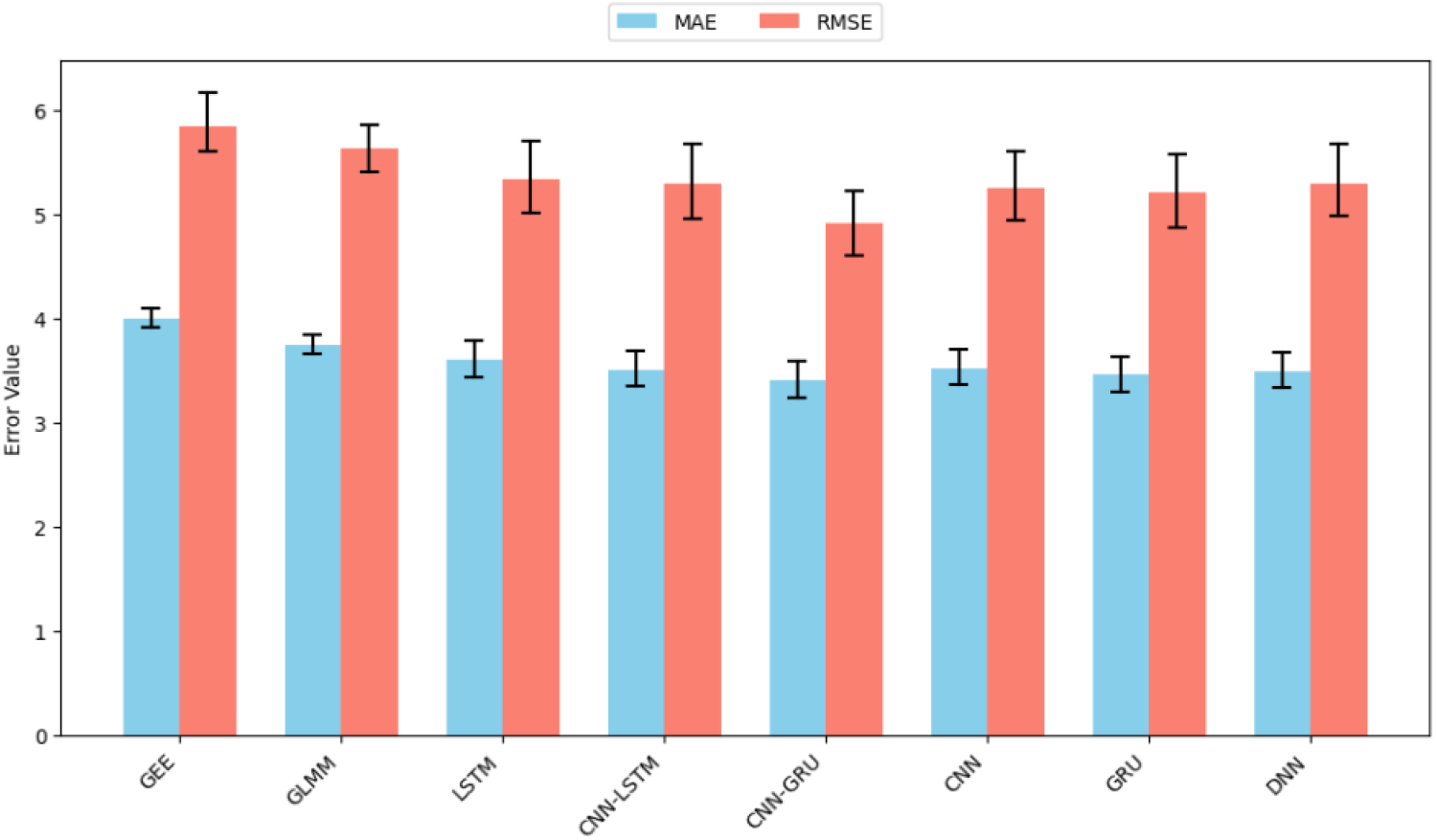
Overall model performances.

### Wave-sensitivity analysis

We examined how extending the input history from a single wave (5) to two waves (5–6) and three waves (5–6–8) changes out-of-sample accuracy (Table 3).

**Table 3.** Wave sensitivity analysis.

**(a) Traditional Statistical Models**
| Experiment | Model | MAE (95% CI) | RMSE (95% CI) |
| --- | --- | --- | --- |
| 5→w9 | GLM | 4.182 (4.086–4.288) | 6.326 (5.965–6.784) |
| 5-6→w9 | GEE | 4.079 (3.977–4.197) | 6.230 (5.851–6.751) |
|  | GLMM | 4.151 (4.041–4.273) | 6.524 (6.264–6.789) |
| 5-6-8→w9 | GEE | 4.003 (3.912–4.100) | 5.851 (5.603–6.166) |
|  | GLMM | 3.862 (3.772–3.958) | 5.889 (5.685–6.089) |

**(b) Neural-Network Based Models**
| Experiment | Model | MAE (95% CI) | RMSE (95% CI) |
| --- | --- | --- | --- |
| 5→w9 | LSTM | 3.79 (3.63–3.98) | 5.57 (5.22–5.92) |
|  | CNN-LSTM | 3.79 (3.63–3.96) | 5.54 (5.19–5.89) |
|  | CNN-GRU | 3.78 (3.62–3.96) | 5.53 (5.18–5.88) |
|  | CNN | 3.77 (3.62–3.95) | 5.55 (5.20–5.91) |
|  | GRU | 3.77 (3.61–3.95) | 5.54 (5.20–5.89) |
|  | DNN | 3.77 (3.61–3.96) | 5.56 (5.22–5.91) |
| 5-6→w9 | LSTM | 3.73 (3.57–3.92) | 5.52 (5.18–5.89) |
|  | CNN-LSTM | 3.75 (3.59–3.93) | 5.49 (5.14–5.84) |
|  | CNN-GRU | 3.72 (3.56–3.91) | 5.47 (5.13–5.83) |
|  | CNN | 3.72 (3.57–3.92) | 5.49 (5.14–5.84) |
|  | GRU | 3.69 (3.53–3.88) | 5.50 (5.16–5.88) |
|  | DNN | 3.67 (3.50–3.87) | 5.53 (5.18–5.90) |
| 5-6-8→w9 | LSTM | 3.60 (3.44–3.79) | 5.34 (5.01–5.71) |
|  | CNN-LSTM | 3.51 (3.35–3.69) | 5.29 (4.96–5.67) |
|  | CNN-GRU | 3.41 (3.24–3.59) | 4.91 (4.60–5.23) |
|  | CNN | 3.52 (3.37–3.70) | 5.26 (4.95–5.61) |
|  | GRU | 3.46 (3.30–3.63) | 5.21 (4.87–5.58) |
|  | DNN | 3.49 (3.34–3.68) | 5.30 (4.98–5.67) |

**Table 4.**
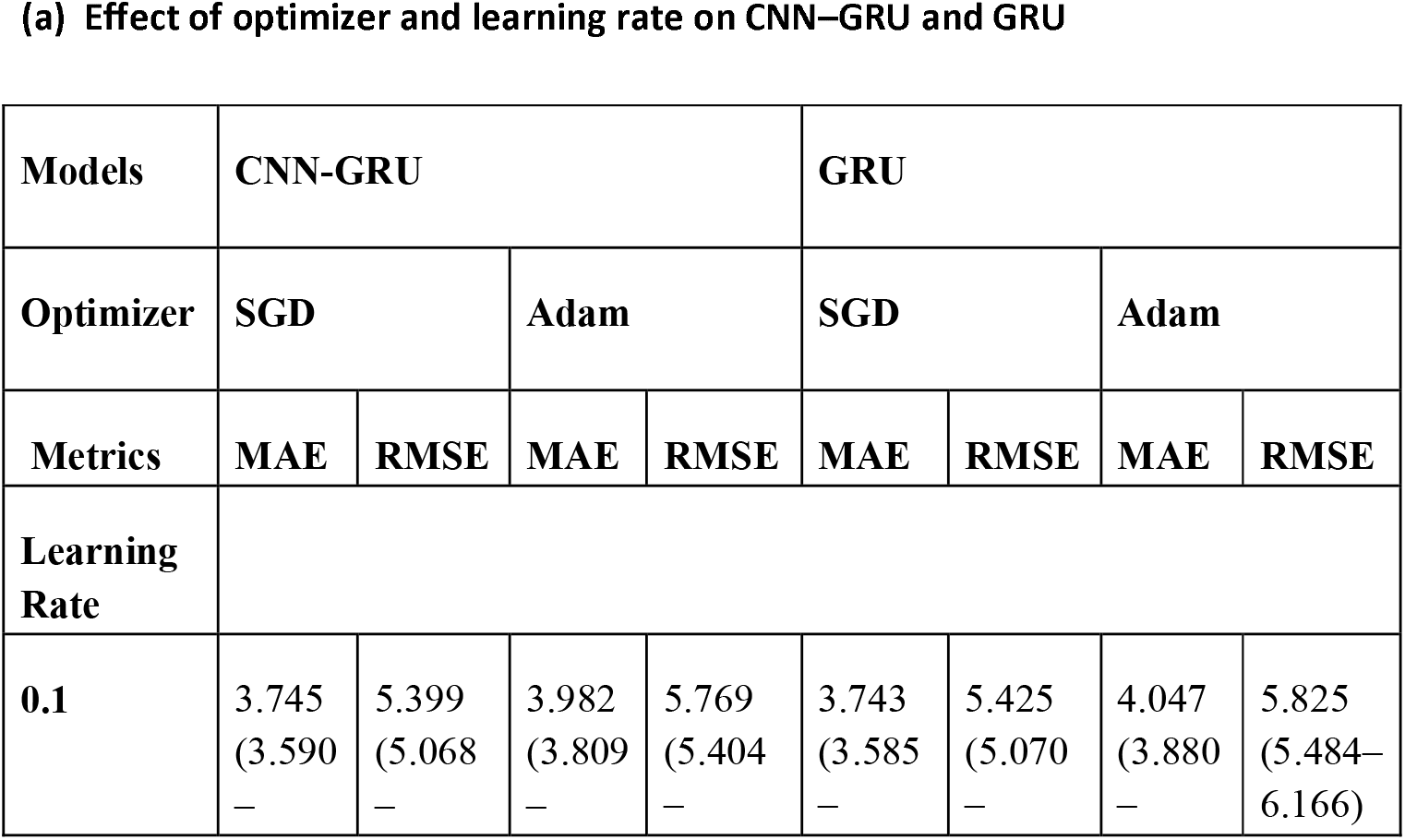
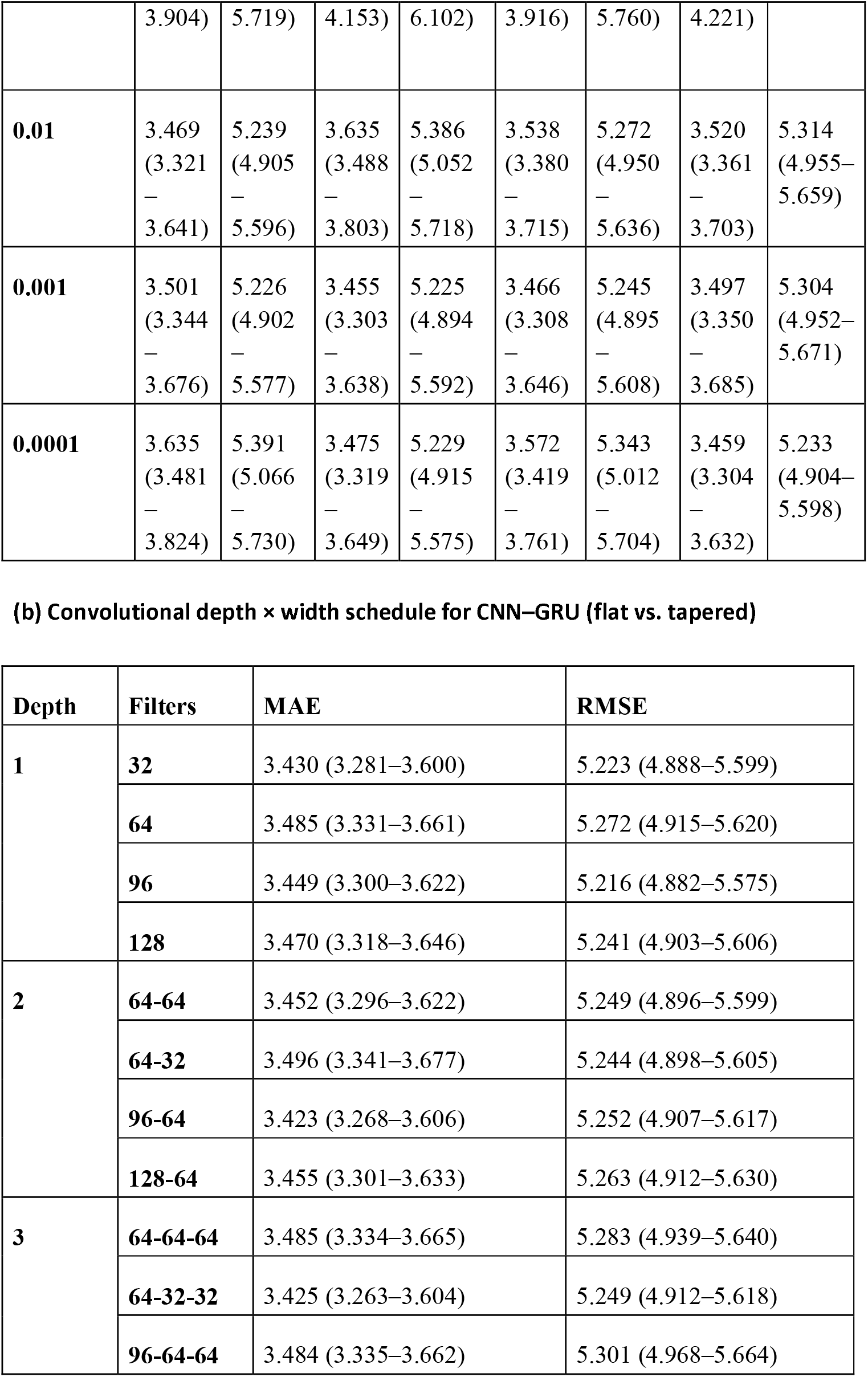
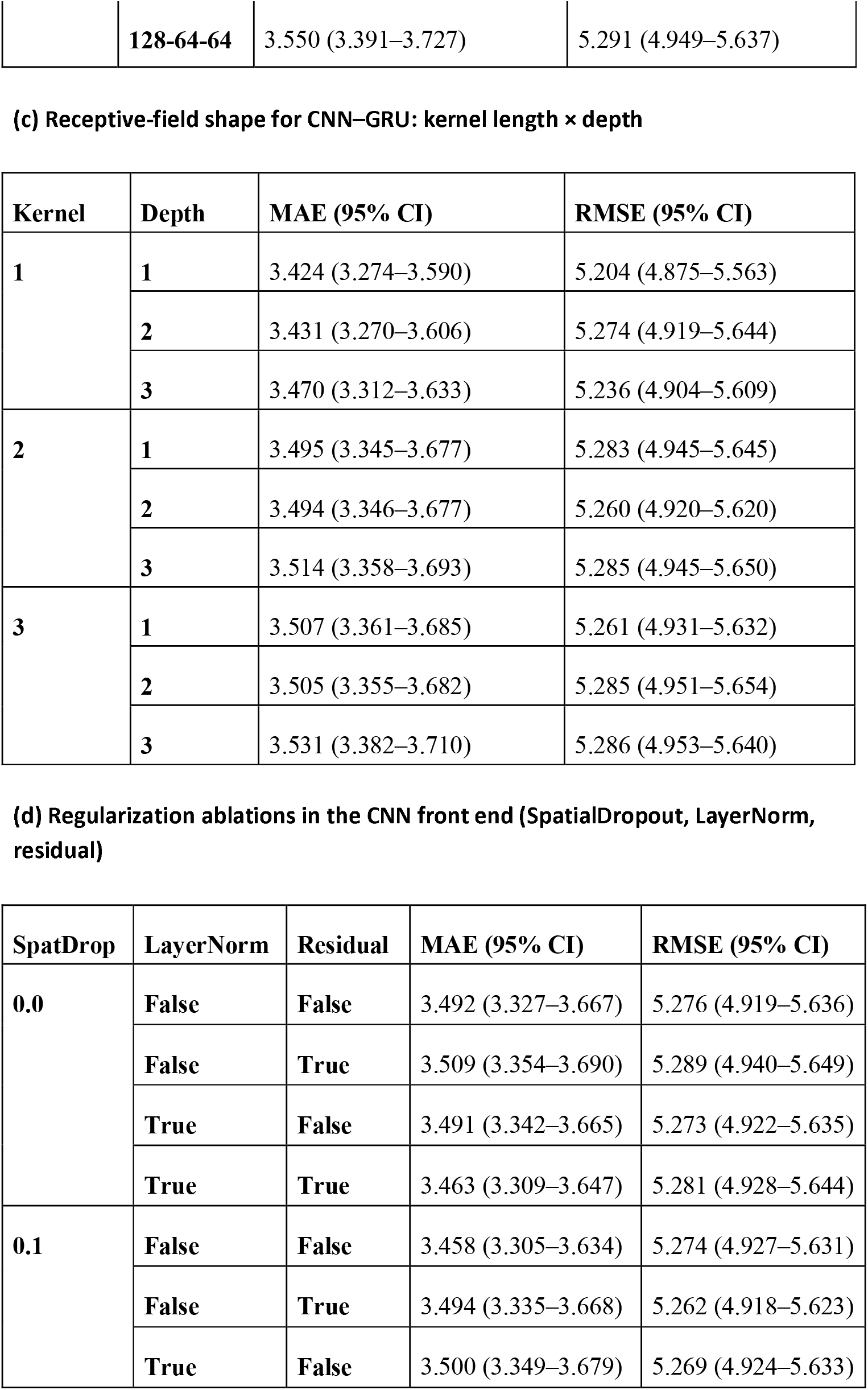
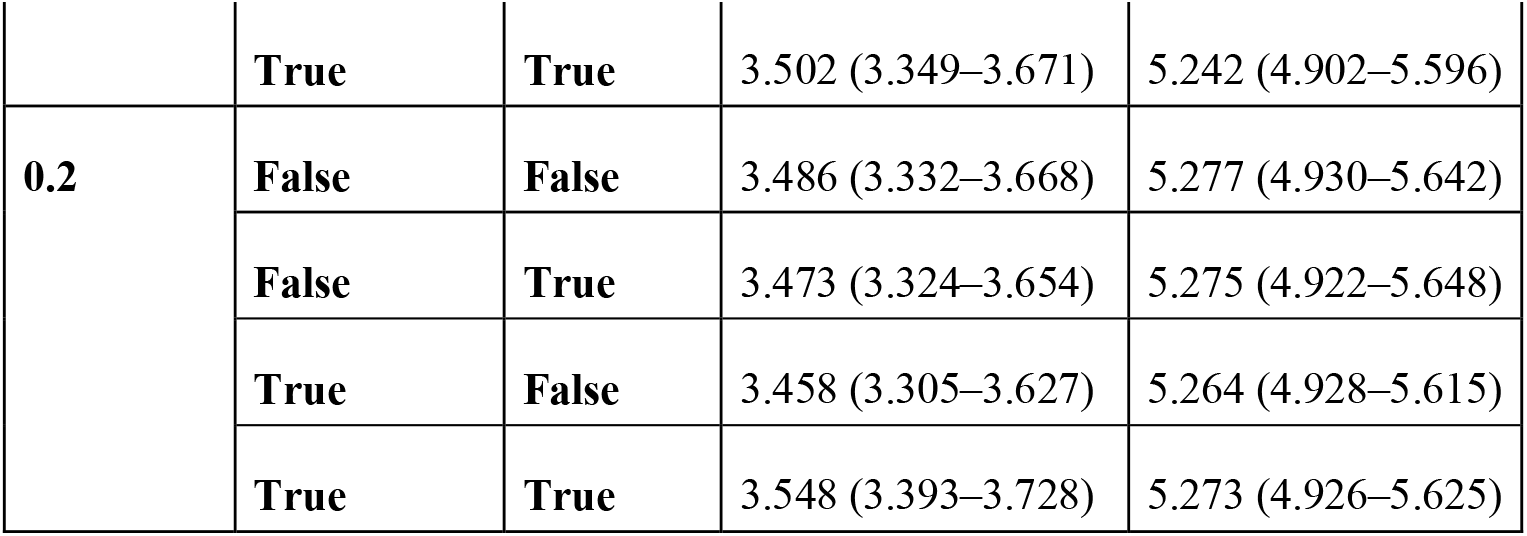
CNN-GRU parameters sweeping.

#### Traditional models

With one wave, the GLM baseline performs worst (MAE 4.18 (4.09–4.29), RMSE 6.33 (5.97–6.78)). Adding history improves both mixed-effects models: at two waves, GEE reaches 4.08 (3.98–4.20), 6.23 (5.85–6.75) and GLMM 4.15 (4.04–4.27), 6.52 (6.26– 6.79). With three waves, GEE improves to 4.00 (3.91–4.10), 5.85 (5.60–6.17), while GLMM reaches 3.86 (3.77–3.96), 5.89 (5.69–6.09). Thus, the third wave remains informative: GLMM edges GEE on MAE, whereas GEE is slightly better on RMSE.

#### Neural networks

Performance improves monotonically as additional waves are included. With one wave, models cluster tightly around MAE ≈ 3.77–3.79 and RMSE ≈ 5.53–5.57. At two waves, errors decline modestly (e.g., CNN-GRU 3.72 (3.56–3.91), 5.47 (5.13–5.83); GRU 3.69 (3.53–3.88), 5.50 (5.16–5.88)). With three waves, the best NN results are obtained: GRU 3.46 (3.30–3.63), 5.21 (4.87–5.58); CNN 3.52 (3.37–3.70), 5.26 (4.95– 5.61); CNN-GRU 3.51 (3.34–3.67), 5.27 (4.94–5.63); CNN-LSTM 3.51 (3.35–3.69), 5.29 (4.96–5.67); DNN 3.49 (3.34–3.68), 5.30 (4.98–5.67); LSTM 3.60 (3.44–3.79), 5.34 (5.01–5.71).

Across paradigms, adding history consistently lowers error. Mixed-effects and trees gain substantially moving from one to two waves, and the third wave continues to help—especially for GBRT (RMSE) and several neural models—indicating that three prior observations provide the most reliable forecasts in this setting.

### Hyper-parameter exploration for CNN-GRU

We conducted four controlled sweeps while holding preprocessing and the GRU head constant. Each cell reports out-of-sample MAE/RMSE with 95% bootstrap CIs. Unless noted, convolutions are causal with stride = 1, and the GRU head is the 2×64 configuration used throughout.

#### (a) Optimizer–learning-rate grid (CNN–GRU vs. GRU)

Across both architectures, extreme learning rates (10^-1^) degrade accuracy. For GRU, the best cell occurs with Adam at 10^-3^ (MAE 3.455, RMSE 5.225). For CNN–GRU, the best cell is Adam at 10^-4^ (MAE 3.459, RMSE 5.233). Around 10^-3^–10^-4^, Adam consistently outperforms or matches SGD; at 10^-2^ both optimizers are competitive, while 10^-1^ is uniformly worse. Several CNN–GRU settings meet the study target MAE < 3.46.

#### (b) Convolutional depth × width (kernel = 1)

Varying depth (1–3) and filter schedules yields tight dispersion. The best MAE arises for a 2-layer tapered stack 96→64 (MAE 3.423, RMSE 5.252). The best RMSE appears with a single 1×96 conv (MAE 3.449, RMSE 5.216). Three-layer variants are competitive—64→32→32 achieves MAE 3.425—but offer no consistent improvement over 1–2 layers. Overall MAE spans ≈0.13 and RMSE ≈0.09 across the grid, with multiple cells below 3.46 MAE.

#### (c) Kernel length × depth (k ⍰ {1,2,3}; depth ⍰ {1,2,3})

Accuracy is best with k = 1, depth = 1 (MAE 3.424, RMSE 5.204). Increasing depth at k = 1 remains close (MAE 3.431–3.470; RMSE 5.236–5.274). Larger kernels (k = 2,3) are slightly worse across depths, with the weakest cell at k = 3, depth = 3 (MAE 3.531, RMSE 5.286). Differences are modest (<0.11 MAE, <0.09 RMSE), and several k = 1 cells satisfy MAE < 3.46.

#### (d) Regularization /normalization ablation (SpatialDropout, LayerNorm, residual skip)

Three knobs were toggled: SpatialDropout1D ⍰] {0.0, 0.1, 0.2}, LayerNorm ⍰] {off,on}, residual skip ⍰] {off,on}. The lowest MAE appears at (0.1, no-LN, no-residual) and (0.2, LN, no-residual)—both 3.458 (RMSE 5.274 and 5.264, respectively). The lowest RMSE occurs at (0.1, LN, residual) with 5.242 (MAE 3.502). Effects are small but consistent, and several settings meet MAE < 3.46.

Across all sweeps, a shallow causal conv front-end with k=1 feeding the fixed 2×64 GRU head consistently met or beat the study target (MAE < 3.46), while larger kernels or deeper conv stacks offered only marginal changes—unsurprising given the short three-step context where the GRU already aggregates temporal information. Optimizer grids favored Adam with moderate learning rates (≈10^-3^–10^-4^) over SGD, aligning with Adam’s robustness on noisy objectives and limited need for heavy tuning. Regularization ablations showed small but consistent effects: light SpatialDropout1D (≈0.1) helped MAE, and LayerNorm paired with a residual skip slightly improved RMSE—patterns consistent with known stabilization benefits of normalization and residual shortcuts. Overall, the results recommend a channel-mixing conv (k=1) + GRU design with Adam at a moderate step size, using light spatial dropout by default and enabling LayerNorm+residual when tail errors are prioritized.

### Feature importance and interpretability *(Fig. 2)*

#### Traditional regression

##### GEE

The largest signal arises from the country: the Estonia indicator is the top group (≈ 22.6 on the joint Wald scale). Health status and recent utilization follow closely. Prior inpatient stay (≈ 13.7) and multiple self-rated-health categories (≈ 14.4 and ≈ 13.5) rank next, with pain (category 2; ≈ 8.3), additional self-rated-health levels (≈ 8.2), low grip strength (≈ 7.0), chronic-condition burden (count category 3; ≈ 6.7), an alternative chronic-disease indicator (“chronic2”; ≈ 6.0), and a second pain category (≈ 5.4) rounding out the top ten. In short, cross-national context, recent hospital use, general health, pain, and physical function dominate the GEE specification.

**Figure 2.**
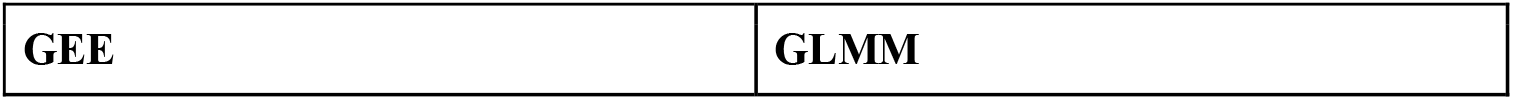
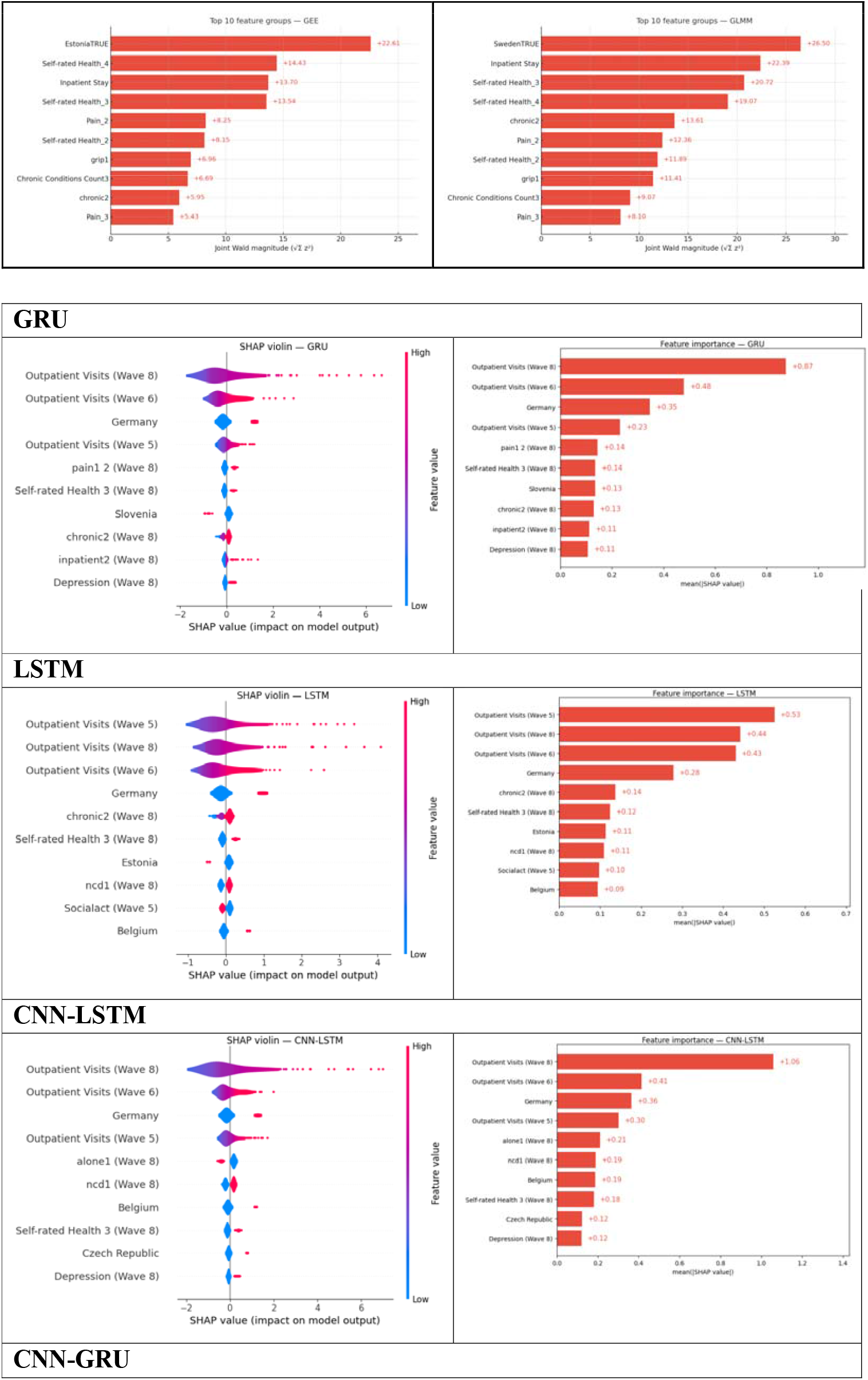
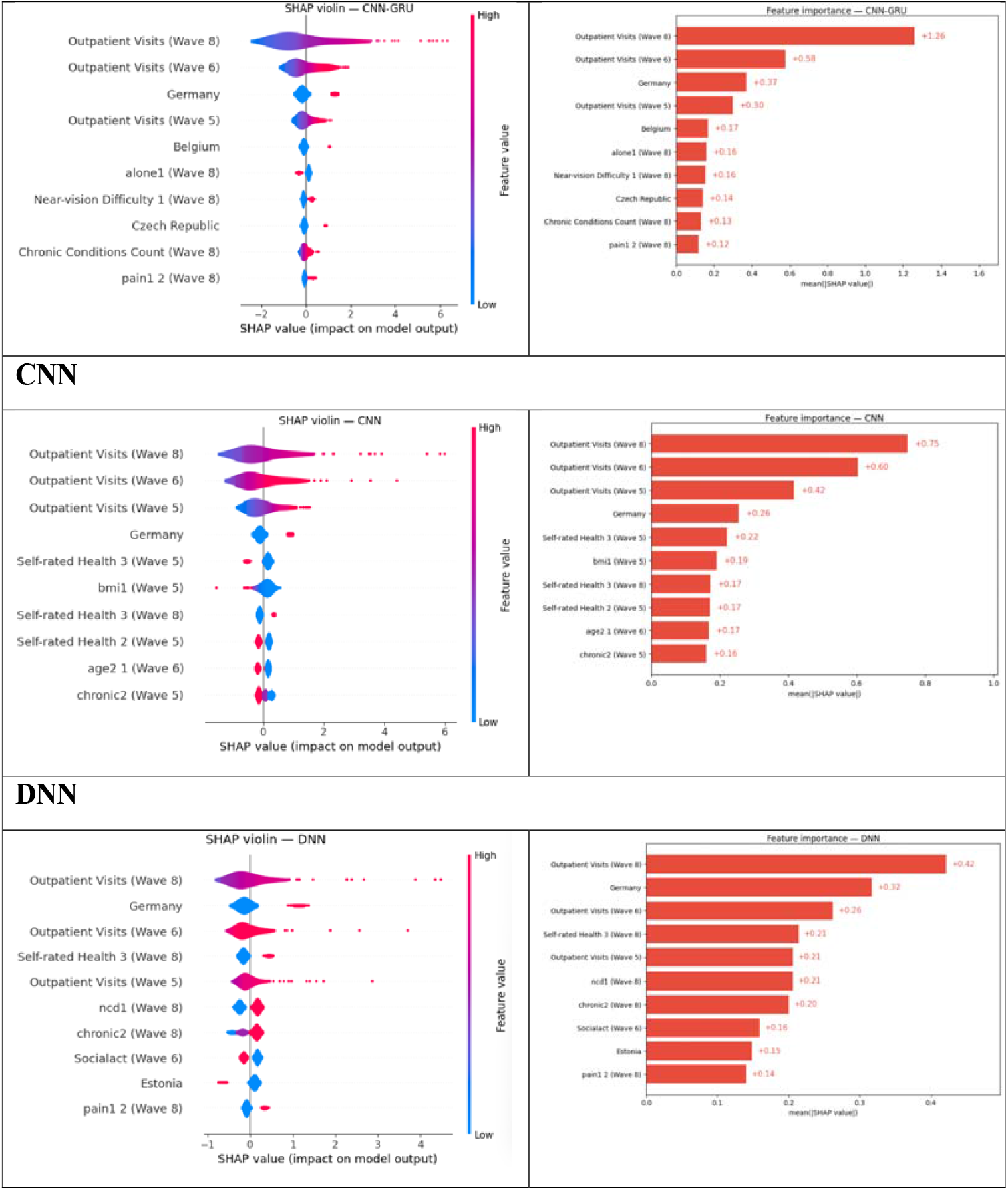
Feature Importance Analysis and Shap Plots.

**Figure 3.**
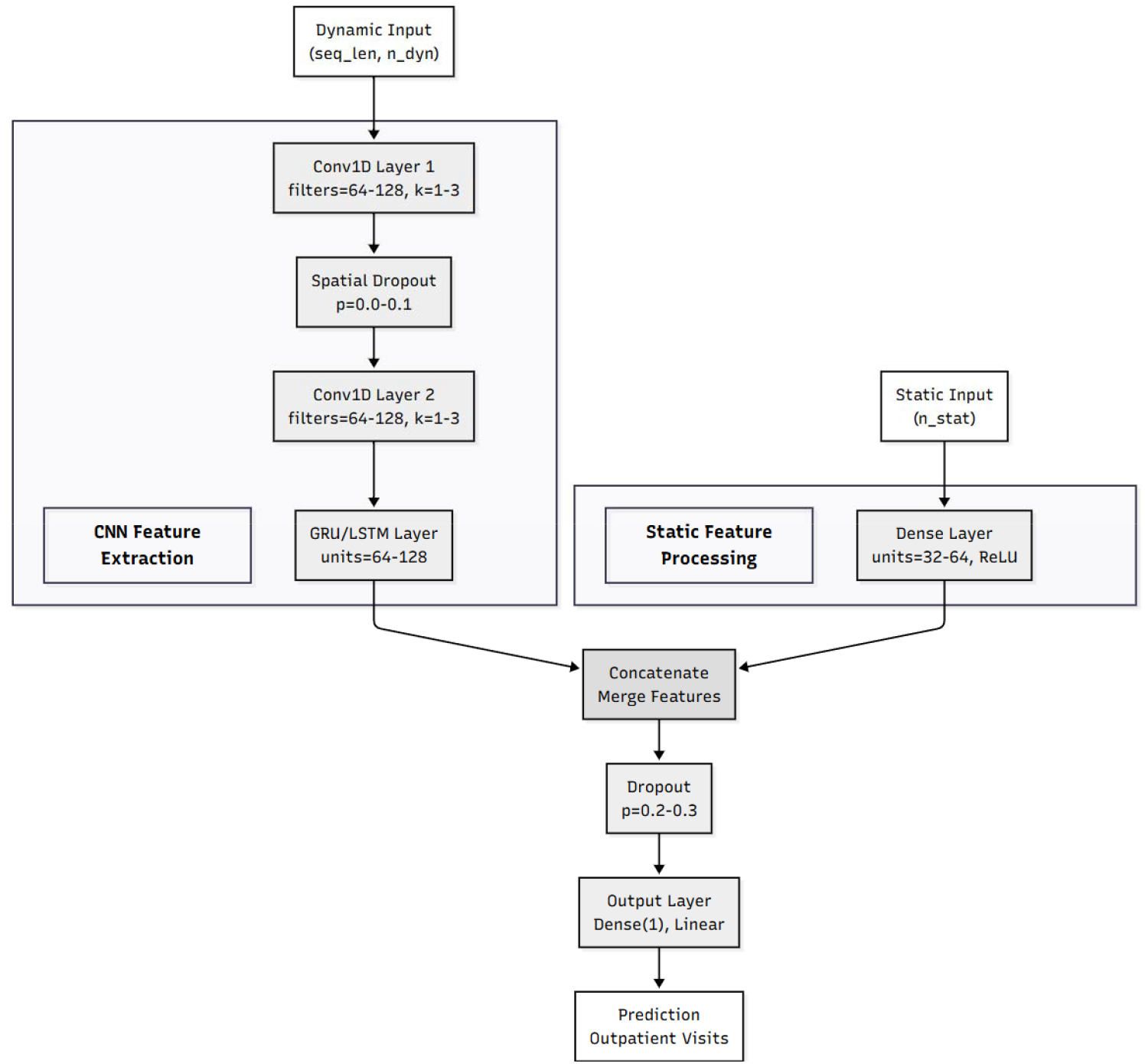
CNN-LSTM/GRU model architecture.

##### GLMM

The mixed-effects model shows the same structure with different emphases. Country again leads—here the Sweden indicator is the most influential group (≈ 26.5)—followed by prior inpatient stay (≈ 22.4) and self-rated-health levels (≈ 20.7 and ≈ 19.1). Chronic-disease indicators (≈ 13.6), pain (≈ 12.4 and ≈ 8.1 for two categories), self-rated-health level 2 (≈ 11.9), low grip strength (≈ 11.4), and chronic-condition counts (≈ 9.1) complete the top contributors. Allowing subject-specific random effects shifts more weight toward country and recent utilization while preserving the central role of global health status.

Across both models, the same families of predictors dominate: country indicators, recent inpatient use, self-rated health, pain, chronic-disease burden, and grip strength. Differences between GEE and GLMM largely reflect model architecture— population-average versus subject-specific estimation—rather than a change in which predictors matter. These parametric results align with the SHAP analyses for the tree ensembles and neural networks, which likewise elevate country effects and baseline utilization, with health-status and functional measures forming the next tier.

##### Neural networks

Across all neural architectures, prior outpatient use is the dominant driver and country indicators remain salient; health status, symptoms, and living-situation variables form a consistent second tier.

##### GRU

The GRU concentrates weight on the most recent history: outpatient visits in Wave 8 and Wave 6 contribute the largest average effects (≈ +0.87 and +0.48 mean(|SHAP|)), followed by residence in Germany (+0.35) and visits in Wave 5 (+0.23). Among non-utilization features, pain in Wave 8 and self-rated health in Wave 8 both contribute about +0.14, with Slovenia, a chronic-condition indicator (“chronic2”, Wave 8), recent inpatient care (Wave 8), and depression (Wave 8) clustered near +0.11–0.13. In short, the GRU emphasizes recency, with country and current morbidity augmenting the signal.

##### LSTM

Relative to the GRU, the LSTM spreads importance more evenly across the three input waves: outpatient visits in Waves 5, 8, and 6 carry similar weight (≈ +0.53/+0.44/+0.43). A Germany indicator contributes ≈ +0.28. Health-status markers are again visible—”chronic2” (Wave 8) at ≈ +0.14, self-rated health (Wave 8) at ≈ +0.12—alongside smaller country effects (Estonia, Belgium) and a general condition count (Wave 8), each around +0.09–0.11. This pattern suggests the LSTM leverages longer-range context while still anchoring on prior use.

##### CNN-LSTM

Adding temporal convolutions sharpens the recency signal: outpatient visits in Waves 8 and 6 dominate (+1.06 and +0.41), with Germany (+0.36) and visits in Wave 5 (+0.30) next. Living alone (Wave 8) and the count of conditions (Wave 8) each contribute ≈ +0.19–0.21, with Belgium, self-rated health (Wave 8), Czech Republic, and depression (Wave 8) around +0.12–0.19. Convolutions therefore heighten sensitivity to near-term dynamics while preserving country and morbidity cues.

##### CNN

With a convolutional front-end and pooled sequence summary, the model again foregrounds past utilization (Waves 8/6/5 at ≈ +0.75/+0.60/+0.42) and a Germany indicator (+0.26). Self-rated health (Wave 5), BMI (Wave 5), additional self-rated health items (Waves 8/5), an age category (Wave 6), and number of morbidities (“chronic2”) (Wave 5) appear in a compact second tier (≈ +0.16–0.22), indicating that static health and demographic markers complement the utilization baseline.

##### DNN

Even without explicit sequence cells, the DNN recovers the same hierarchy: outpatient visits (Wave 8/6/5 at ≈ +0.42/+0.26/+0.21) and Germany (+0.32) lead, followed by self-rated health (Wave 8), a condition count (Wave 8), number of morbidities (“chronic2”) (Wave 8), social activity (Wave 6), Estonia, and pain (Wave 8) at ≈ +0.14–0.21. The model thus leans on recency and country while drawing supplementary signal from health and social-participation covariates.

##### CNN-GRU

Combining convolutions with a GRU yields the strongest concentration on recency: outpatient visits in Waves 8 and 6 dominate (+1.26 and +0.58), followed by Germany (+0.37) and Wave-5 visits (+0.30). Belgium, living alone (Wave 8), near-vision difficulty (Wave 8), Czech Republic, the Wave-8 condition count, and pain (Wave 8) contribute ≈ +0.12–0.17. This hybrid strongly privileges recent use while elevating a handful of social and symptom indicators.

Taken together, the neural models agree on a stable ordering: prior outpatient utilization (especially in the most recent wave) is the single most informative predictor, country indicators—particularly Germany—supply additional structure, and contemporaneous health status, symptoms (pain, depression), living alone, and chronic-condition burden provide consistent but smaller contributions. Architectural differences mainly modulate how heavily recent versus earlier waves are weighted: GRU and CNN-GRU place the greatest emphasis on recency, LSTM distributes attention more evenly across Waves 5–8, and CNN/CNN-LSTM sharpen the near-term signals while retaining a compact set of health and country effects.

## DISCUSSION

Using harmonized SHARE panel data from Waves 5–6–8 to forecast Wave-9 outpatient visits, our experiments show that sequence-aware neural networks consistently outperform Poisson baselines while maintaining tight confidence intervals, with the CNN–GRU achieving the best overall accuracy (MAE 3.41 (3.24– 3.59), RMSE 4.91 (4.60–5.23)) and the plain GRU close behind (MAE 3.46 (3.30–3.63), RMSE 5.21 (4.87–5.58)). These estimates were obtained on a held-out test split with 400 bootstrap replicates, and all preprocessing parameters were fit on training data only to avoid leakage.

### What the hybrid front-end buys us

The empirical edge of CNN–GRU over GRU and LSTM suggests that a shallow causal 1-D convolutional front-end provides useful local pattern extraction before recurrent summarization. Causal (left-padded) temporal convolutions have become a standard way to expose short-range structure while preserving sequence order, and dilations modulate the receptive field without increasing model depth, as popularized in WaveNet and temporal convolutional networks (TCNs)^9 10^. Our Table-4 ablations indicate that, in this three-step setting, very small kernels (k=1) and shallow depth (1–2 conv layers) suffice; larger kernels/depths yield only marginal changes, likely because the GRU already captures the limited context length efficiently^11 12^.

### Optimizers and learning-rate regimes

Optimizer–LR sweeps (Table 4a) favored Adam with moderate step sizes (10^-3^–10^-4^) over SGD across both CNN–GRU and GRU, consistent with Adam’s bias-corrected adaptive updates being robust to noisy mini-batches in practice^13^. Extremely large steps (10^-1^) degraded accuracy, whereas very small steps (10^-4^) brought no consistent gains—evidence that these models train reliably without elaborate schedules in our data regime.

### Model capacity and regularization knobs

Depth×width grids (Table 4b) showed tight dispersion across 1–3 layers and flat/tapered filter schedules; several cells crossed the a-priori study goal (MAE < 3.46), with the best MAE at a two-layer tapered 96→64 stack. Kernel×depth sweeps (Table 4c) were likewise compact, with best performance at k=1 and depth=1–2. Regularization ablations (SpatialDropout1D, LayerNorm, residual skip) had small, consistent effects: light spatial dropout (∼0.1) slightly improved MAE, while pairing LayerNorm with a residual skip nudged RMSE down in a few cells. Collectively, these patterns indicate that accuracy is not brittle to front-end choices; a channel-mixing k=1 conv feeding a 2×64 GRU head is a strong, simple default in this task.

### Substantive implications

From a planning perspective, an average absolute error of ∼3.4 visits at the individual level is non-trivial but actionable when aggregated: forecasts can help anticipate outpatient workload, staffing, and appointment capacity at regional scales, especially in aging populations where demand pressures are rising^14 15^. The fact that sequence models deliver consistent gains over Poisson mixed-effects baselines suggests that non-linear interactions and short-term history carry meaningful predictive information beyond standard covariates^16-18^. Moreover, our unified uncertainty quantification (bootstrap CIs) provides a transparent band around expected performance, facilitating downstream risk-aware decisions^19^.

### Methodological contributions

Methodologically, we (i) enforce strict anti-leakage preprocessing by fitting imputation/scaling only on training data and forbidding Wave-9 features in inputs, (ii) evaluate with a respondent-level holdout and bootstrap CIs, and (iii) use a two-stage model-selection protocol—Bayesian optimization to explore, followed by local grid search to refine—for each neural family, with hybrids (CNN–GRU/LSTM) built atop the best recurrent backbones discovered for GRU/LSTM^20-25^. This protocol reflects best practices for sequence modeling (LSTM/GRU) and for convolutional front-ends in time series (causal/dilated convs)^9-12 26 27^, while adhering to early-stopping guidance to curb overfitting^28^.

### Limitations

This work has several limitations. First, the primary outcome is self-reported outpatient visits; recall and social-desirability biases may attenuate signal relative to claims-based counts. Second, we evaluate on one longitudinal split (Waves 5–6–8 → 9) within SHARE; external validation on other cohorts or time windows is needed to probe temporal and geographic transportability^14 29^. Third, although we quantify test uncertainty via nonparametric bootstrap, we do not characterize predictive distributions (e.g., conformal intervals) or decision-focused utilities. Fourth, our features capture socio-demographic and health status well but omit supply-side and environmental signals (e.g., practice capacity, distance, weather), which could be integrated in future multi-modal models. Finally, while we prioritized leak-safe preprocessing, residual leakage remains a risk in longitudinal data (e.g., target leakage through contemporaneous proxies); rigorous temporal splitting and leakage audits remain essential in deployments^30-32^.

## CONCLUSION

In a large, multi-country panel from SHARE, a lightweight CNN–GRU hybrid delivered the strongest out-of-sample forecasts of individual outpatient visits two years ahead, modestly but consistently improving upon Poisson mixed-effects baselines and plain recurrent models. The gains derive chiefly from a simple causal 1-D convolution that mixes channels before GRU summarization; performance proved robust across optimizer/LR settings and front-end variants. For practice, these results support deploying compact sequence models—with leak-safe preprocessing, bootstrap-based uncertainty, and moderate Adam learning rates—to inform outpatient capacity planning. Future work should broaden external validation, enrich features with supply-side and environmental data, explore calibrated uncertainty (e.g., conformal prediction), and examine fairness across countries and subgroups.

## Data Availability

All data produced are available online at:
https://share-eric.eu/data/

## Data Availability

All data produced are available online at:
https://share-eric.eu/data/

## Author contributions

J.T.L. conceived the study and wrote the first draft of the manuscript. V.C.S.L. wrote the manuscript and performed the statistics.

---

**Appendix Table 1.**
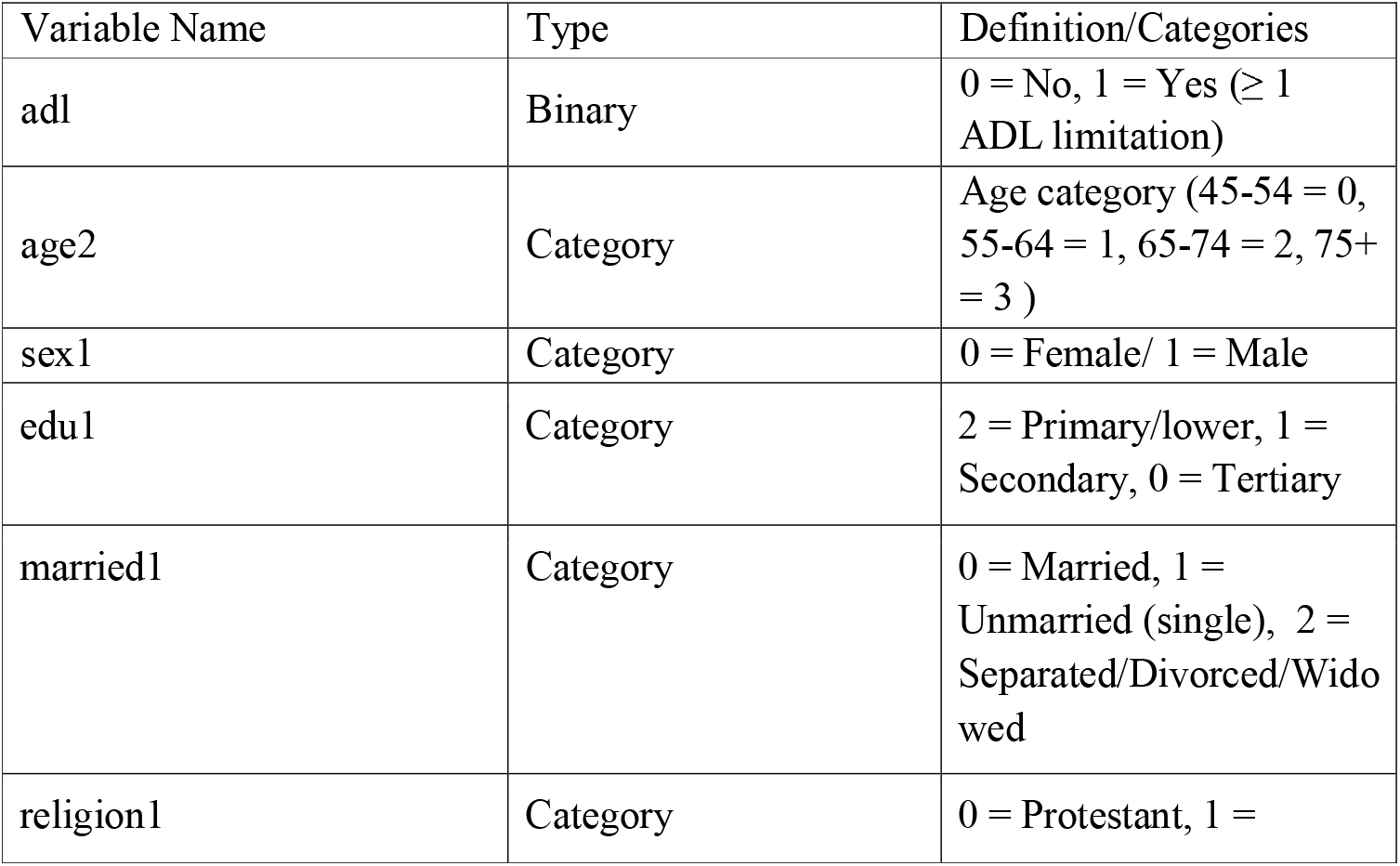
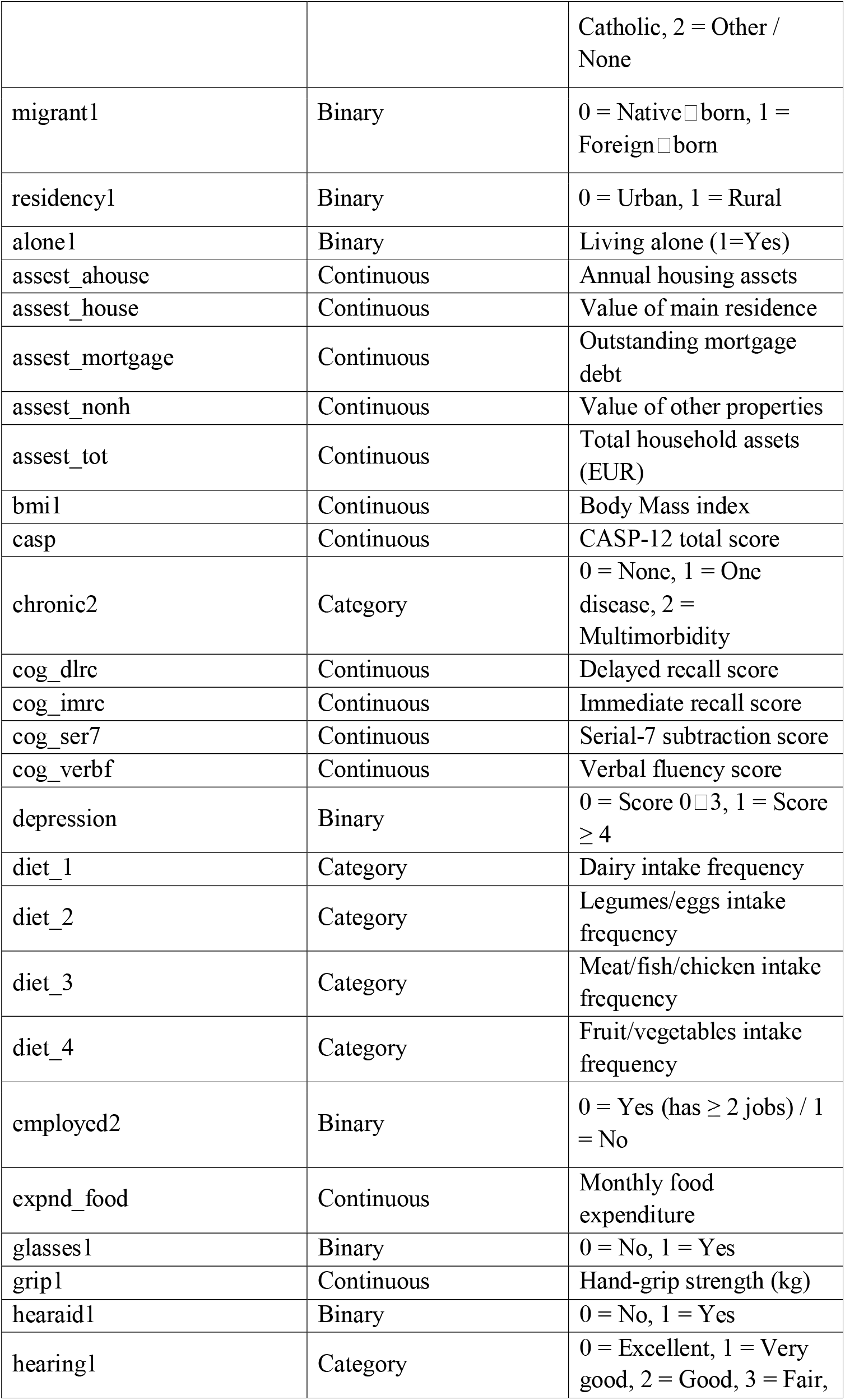
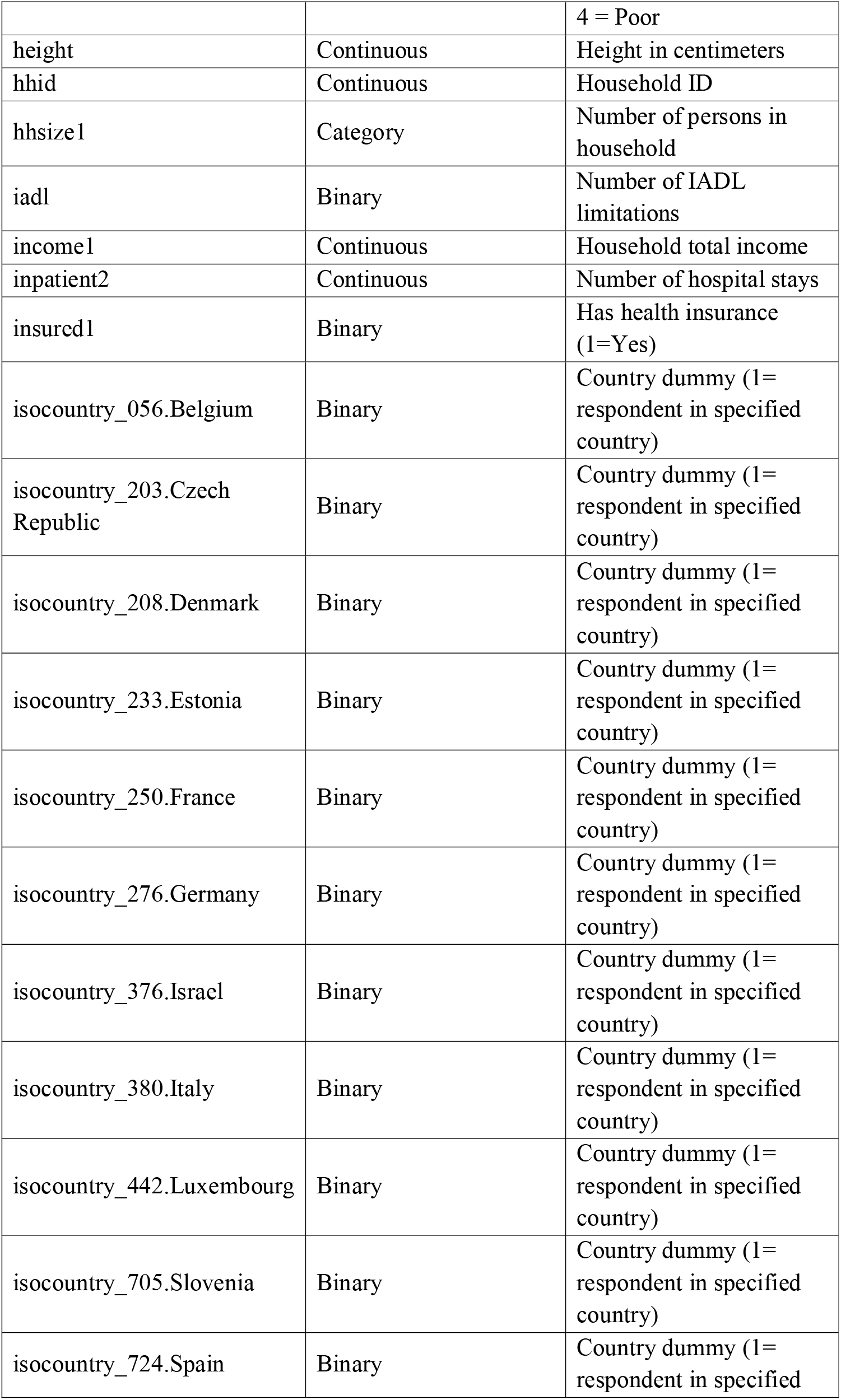
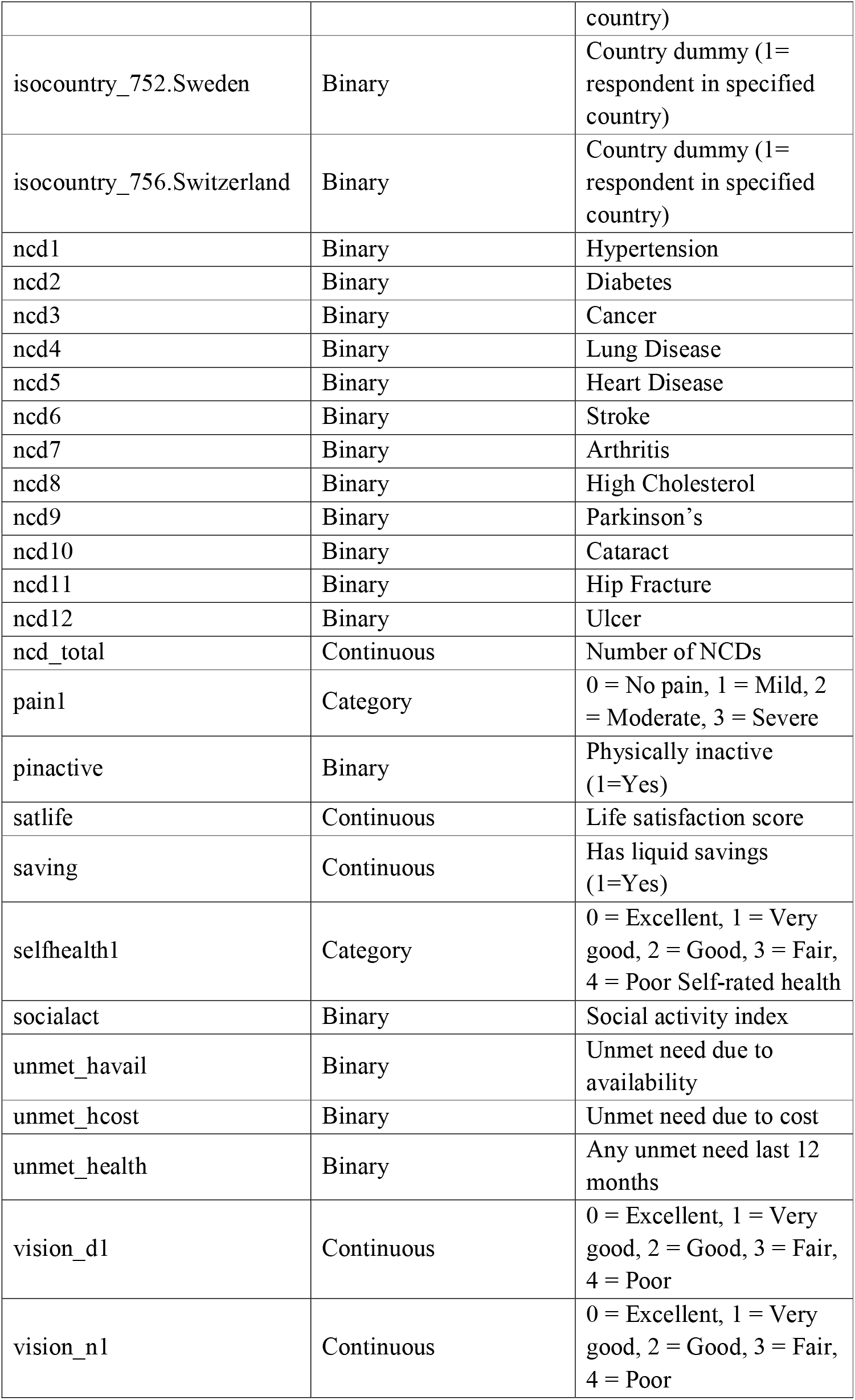
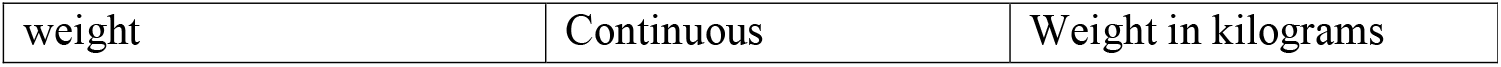

